# Assessment of the accessibility of gyms for people with disability: A case study in New Zealand

**DOI:** 10.64898/2026.08.02.26359531

**Authors:** Claire Flemmer, Mikael Boulic

## Abstract

**Background:** Access to exercise facilities is particularly important for the health, wellbeing, and social participation of people with disability. However, many facilities remain difficult for them to access.

**Methods:** This research used a mixed-method case study of twenty-three gyms in New Zealand. Quantitative accessibility audits based on the requirements of NZS4121:2001 were used to assess the compliance of accessible parking, entrances, interiors and bathrooms. Interviews with staff or managers at each gym provided qualitative data on operational practice and awareness of inclusivity.

**Results:** The most accessible aspects include parking spaces, footpaths, entrance doors and bathrooms with features such as clear signage, well-designed ramps, and evacuation protocols demonstrating best practice inclusivity. Improvements in service counters, lifts, tactile and auditory communication are needed. Community gyms and university gyms are the most inclusive and their staff have the most comprehensive experience in interacting with people with different disabilities. Small private gym owners know little about accessibility barriers and financial constraints limit their ability to improve access features.

**Conclusion:** The main recommendations for improving gym accessibility for people of all abilities include strategies to facilitate their joining gyms and strategies to support them as long-term gym members. The involvement of healthcare professionals in prescribing regular exercise and monitoring the health effects, coupled with subsidised gym membership will make it easier for people with disability to join gyms. More extensive staff training, targeted exercise programs, inclusive signage and communication and regular maintenance of the facility will improve the ongoing experience. Participating in regular exercise and social environments will help improve the physical and psychological health of people with disability.

## 1. Introduction

The proportion of people with disability (PWD) is estimated at 16% of the global population (WHO, 2023) and 17% of New Zealand’s population (Household Disability Survey, 2023). On average, PWD are more prone to secondary health conditions, such as obesity (Anderson et al., 2013), diabetes (Jung et al., 2021) and cardiovascular diseases (Jerome et al., 2024) and are also more likely to have psychological health conditions such as depression and anxiety (Sharon-David et al., 2021). Physical activity benefits both physical and psychological health and for PWD the benefits can lead to greater independence and improved quality of life (Rimmer et al., 2004). For this reason, exercise is frequently recommended (Verschuren et al., 2016; Halabchi et al., 2017; Leathem et al., 2021; Sezer et al.,2015). However, PWD participation in indoor exercise, recreation and sporting facilities (referred to here as the generic term “gyms”) is low. Studies on people using wheelchair have shown that fewer than half do any physical activity and only 15% of them exercise enough to see health benefits (Morgan et al., 2024). Similarly low participation rates have been found for people with other types of disabilities (Sharon-David et al., 2021; Lesch et al., 2025). Many countries have acknowledged this issue and have developed guidelines to address it. Table 1 provides a sample of recent guidelines.

**Table 1.** Recent global guidelines for improving gym accessibility and inclusivity for PWD.

| Country | Organisation (Year) | Description |
| --- | --- | --- |
| New Zealand | Sport New Zealand (2023) | National Indoor Active Recreation and Sport Facilities Strategy. |
| Australia | Sport and Recreation Victoria (2024) | Design for Everyone Guide - A guide to Sport and Recreation Settings. |
| Australia | Australian Sporting Alliance for People with a Disability ASAPD (2025) | Disability Sports – Community Facility Guidelines. |
| United Kingdom | Sport England (2024) | Accessible and Inclusive Sports Facilities (AISF). |
| Ireland | Disability Sport Northern Ireland (2022) | Accessible Sports Facilities Design Guidelines (2022). |

One of the main barriers to PWD using gyms is poor accessibility. Article 9 of the United Nations Convention on the Rights of Persons with Disabilities (CRPD) states that signatory countries shall take the appropriate measures “*to enable persons with disabilities to live independently and participate fully in all aspects of life*” (United Nations, 2006). This requirement encompasses the built environment, including gyms.

Researchers have found that there are both structural and social barriers to inclusive gym accessibility (Rolfe et al., 2016). They have investigated the barriers from the perspective of different stakeholders including gym staff and gym users (both with and without disabilities). They have also developed a range of tools to measure accessibility. These aspects are reviewed below.

### 1.1 The perspectives of gym staff on inclusive accessibility

Practices for social support in gyms include orientation, supervision, specialist input, logistical support, motivation, peer support and social activities (McKenzie et al., 2025). They are used to support both new members and to support ongoing participation. However, staff in government-funded gyms report that little or nothing is done to encourage PWD to join the gym. Instead, the initial contact occurs when the PWD themself or a school or supporting organisation enquires about using the gym facilities. Once the PWD joins, the social support varies widely across the gyms but is primarily provided by disability groups rather than by the gym staff. The gyms do not offer educational support to inform PWD on the benefits of physical activity. Activities within the gyms consist of disability-specific exercise groups (supervised by disability organisations) which are mostly closed to other members as well as some informal social activities for everyone. PWD do not often use personal trainers. Instead, the gym provides support to PWD either through a key engagement coordinator or through a volunteer staff “champion” who wants to provide PWD support. Most gym staff receive basic disability awareness training but would like more detailed training on how to interact with people with different types of disabilities and how to provide their exercise needs safely. Cunningham et al. (2023) report that staff who interact regularly with PWD have mostly positive interactions and increased confidence in meeting the needs of PWD. Government and management policy/funding dictates the priorities for social inclusion initiatives. According to McKenzie et al. (2025) and Kennedy et al. (2022), carer and physiotherapist support vary across the gyms; some facilities allow free access to carers holding companion support cards. External physiotherapists are not usually allowed into the gym without a formal arrangement. In both cases, there are issues regarding who is responsible for the safety of the PWD.

### 1.2 Perspectives of PWD on accessing gyms

Research by Flemmer & McIntosh (2025) on twenty-nine different types of public venues in New Zealand found that gyms are perceived by PWD to be difficult to access, with only half of PWD ever going to a gym. This is not necessarily an accessibility issue; lack of motivation is a contributing factor (Georgiou & Kaprinis, 2022). Global studies show PWD may feel intimidated, embarrassed and unwelcome in gyms which are stereotypically regarded as environments for young, fit, muscular people (Richardson et al., 2017a, Rivera et al., 2024). PWD may also feel anxious about their safety while exercising, their risk of experiencing pain, their ability to understand how to use the exercise equipment, and whether they are physically capable of using it (for example, because they are much weaker than other users or need more space around the equipment to manoeuvre a wheelchair). Additionally, they may be worried about negative reactions from other gym members (Sharon-David et al., 2021; Calder et al., 2018; Martin Ginis et al., 2021). PWD have considerably less “gymtimidation” in gyms that are designed on the principles of Universal Design with inclusive signage; clear multi-mode (braille, audio, pictorial and visual) communication systems; adaptable gym equipment; adequate space around equipment; accessible entrances without stairs; and services such as a welcoming atmosphere, support for carers and informative websites (Mitchell et al., 2025 a,b; O’Sullivan et al., 2020). Brown et al. (2021) and Clouse et al. (2020) report that the needs of people with sensory and cognitive impairments are under researched. Finally, the involvement of PWD in evaluating the usability of gyms is essential (Mosca & Capolongo, 2020).

### 1.3 Evaluating gym accessibility

A challenge in advancing inclusive fitness environments is that accessibility is often discussed in theoretical terms, while practical evaluation requires measurable criteria. A key contribution in this area has been the development of structured instruments designed to assess accessibility in fitness and recreation settings. Rimmer et al. (2004) developed and validated AIMFREE (Accessibility Instruments Measuring Fitness and Recreation Environments), providing a systematic method to evaluate facility accessibility. The AIMFREE framework has supported later work examining health club accessibility for people with mobility and visual impairments (Rimmer et al., 2005a,b) and broader assessments indicating that fitness facilities still lack accessibility for PWD (Rimmer et al., 2017). In addition to measurement tools, conceptual frameworks have been proposed to guide improvement. Riley et al. (2008) presented a conceptual framework for improving accessibility of fitness and recreation facilities for PWD, emphasising that inclusive participation depends on both environmental and organisational conditions. Empirical audits also suggest that compliance with accessibility legislation and standards is inconsistent. For example, Stoelzle and Sames (2014) assessed the compliance of American metropolitan gyms with the Disabilities Act highlighting persistent gaps between the intent of the policy and its implementation at the facility-level.

New Zealand has accessibility legislation and standards intended to support equitable access to public and private facilities. The New Zealand Standard NZS 4121:2001 - Design for Access and Mobility provides measurable criteria for the built environment. This includes specific requirements for accessible routes, gradients, door widths, circulation, lifts, and sanitary facilities (Standards New Zealand, 2001). However, the existence of policy does not guarantee consistent implementation. Despite comprehensive regulatory frameworks (such as New Zealand’s Building Act and associated standards), there remain gaps in compliance for public building accessibility and uneven enforcement of accessibility expectations (Flemmer & McIntosh, 2024).

Compared with public spaces such as cultural venues and educational institutions, gyms are relatively under-researched in relation to accessibility, despite their role as community spaces supporting wellbeing and social participation (Dawson, 2017).

Despite growing international evidence that fitness and recreation facilities often remain inaccessible for people with disability, there are several gaps that warrant attention in New Zealand. First, there is limited published research examining gym accessibility across different facility types (such as community facilities, large chains, and small private gyms), and the New Zealand context is underexplored. Second, studies often focus primarily on physical features (e.g., entrances, door widths, and parking), while operational factors such as staff training, awareness of standards, and emergency procedures may strongly influence real-world accessibility. Third, there remains a need for mixed methods approaches that integrate measurable audits with qualitative perspectives to support actionable improvements.

Accordingly, the aim of this study was to assess the accessibility of different types of gyms in New Zealand for people with disability, and to identify barriers and opportunities for improvement across both built-environment and service dimensions. This was achieved through structured accessibility audits aligned with NZS 4121:2001 requirements and staff/manager interviews to capture operational insights. Four types of gyms were considered, namely government-subsidised (not-for-profit) community facilities, large chain gyms, gyms on university campuses and private/independently owned gyms.

## 2. Methods

This research used a mixed-methods case study design, combining quantitative accessibility audits with qualitative staff/manager interviews. A mixed-methods approach was selected because physical compliance measures alone do not fully capture usability in practice, while interviews alone cannot objectively verify whether facilities meet measurable accessibility requirements.

The study focused on fitness facilities in New Zealand, with case study data collected across multiple sites and facility types. Facilities were identified using online searches (e.g., Google Maps and gym directories), and facility managers were contacted by phone, email, or in person to request participation and schedule site visits. Audit data was collected for:

- Eleven gyms in Auckland comprising four government-managed public leisure centres, four large-scale chain gyms, one university gym and two independent (privately-owned) gyms.
- Twelve gyms in Christchurch comprising four government-managed public leisure centres, three large-scale chain gyms, three university gyms and two independent gyms.

Physical accessibility was assessed using structured checklists aligned with NZS 4121:2001 (Design for Access and Mobility). Audited areas included accessible parking, main entrance, interiors and accessible bathrooms, with sub-features shown in Table 2.

**Table 2.**
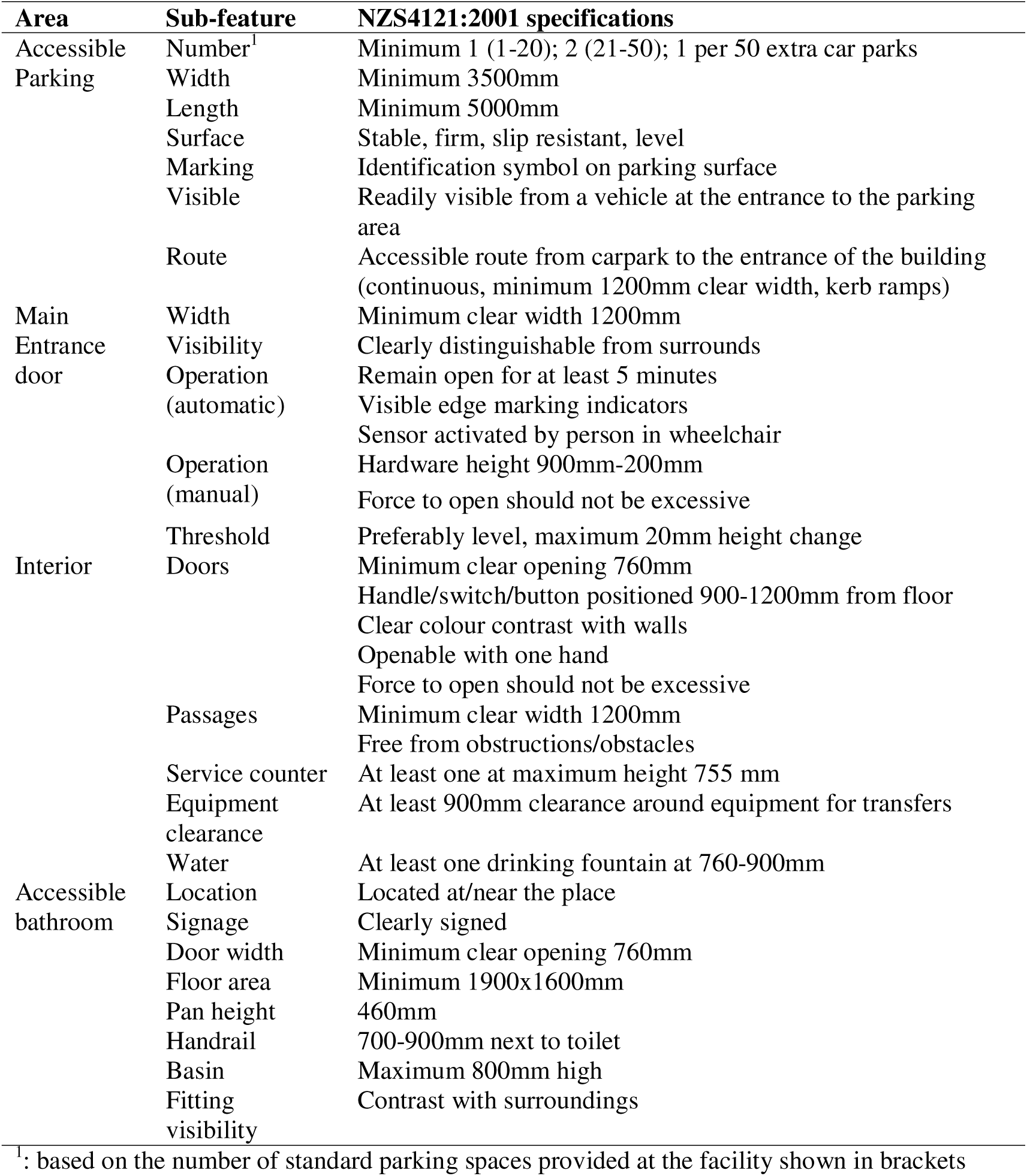
Checklist of areas and sub-features with NZS4121:2001 specifications.

Measurements were collected using standard tools such as measuring tapes and, where appropriate, mobile applications for gradient assessment. Each feature was rated as pass (compliant) or fail (non-compliant) based on whether it met the standard. Items not applicable or not present were recorded separately and excluded from feature-specific compliance calculations. Audit data were summarised using descriptive statistics, with compliance outcomes presented as percentages.

Qualitative data were collected through interviews with eleven facility staff and twelve managers/owners. The interviews included both open-ended questions and questions requiring a 5-point Likert scale response. Interviews explored awareness of accessibility standards, operational challenges in supporting gym users with disability, training and preparedness, emergency procedures, and suggested improvements. Interviews were audio-recorded with consent and transcribed for analysis.

Interview transcripts were analysed using thematic analysis to identify patterns in staff perceptions of accessibility and service delivery barriers. Findings were triangulated by cross-referencing audit outcomes with interview insights to identify areas where physical accessibility issues aligned with operational concerns.

Ethical procedures included obtaining informed consent from interview participants, protecting confidentiality through anonymisation, and conducting site visits in a manner that minimised disruption to facility operations. Low-risk ethics applications (ID 4000029402, 4000029763, 4000029937 granted on 29/08/2024, 17/9/2024, 29/11/24 respectively) were recorded by the university’s human ethics committee.

## 3. Results

The results are presented in two sub-sections, namely, the results of the gym audits (for accessible parking, main entrance, internal features and accessible bathrooms) and the results from the interviews with gym personnel.

### 3.1 Accessibility audit findings

#### 3.1.1 Accessible parking

Seven aspects of accessible parking were considered, namely the number provided, width, length, surface quality, surface marking, visibility from the parking entrance and provision of an accessible route to the gym entrance. Each aspect was judged as a “Pass” if it met or exceeded the NZS4121: 2001 recommendation. Table 3 summarises the results.

**Table 3.**
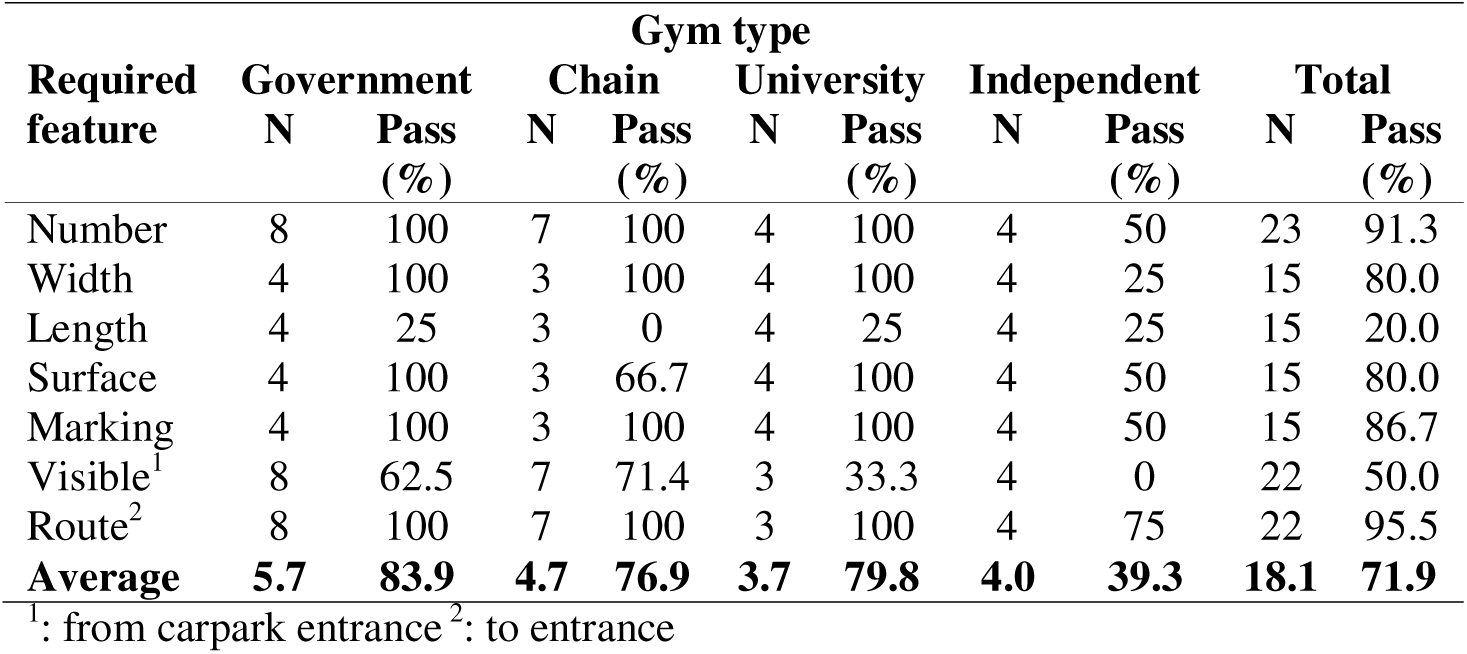
Audit results for accessible parking features in different types of gyms.

| Required feature | Gym type |  |  |  |  |  |  |  |  |  |
| --- | --- | --- | --- | --- | --- | --- | --- | --- | --- | --- |
|  | Government |  | Chain |  | University |  | Independent |  | Total |  |
|  | N | Pass (%) | N | Pass (%) | N | Pass (%) | N | Pass (%) | N | Pass (%) |
| Number | 8 | 100 | 7 | 100 | 4 | 100 | 4 | 50 | 23 | 91.3 |
| Width | 4 | 100 | 3 | 100 | 4 | 100 | 4 | 25 | 15 | 80.0 |
| Length | 4 | 25 | 3 | 0 | 4 | 25 | 4 | 25 | 15 | 20.0 |
| Surface | 4 | 100 | 3 | 66.7 | 4 | 100 | 4 | 50 | 15 | 80.0 |
| Marking | 4 | 100 | 3 | 100 | 4 | 100 | 4 | 50 | 15 | 86.7 |
| Visible<sup>1</sup> | 8 | 62.5 | 7 | 71.4 | 3 | 33.3 | 4 | 0 | 22 | 50.0 |
| Route<sup>2</sup> | 8 | 100 | 7 | 100 | 3 | 100 | 4 | 75 | 22 | 95.5 |
| **Average** | **5.7** | **83.9** | **4.7** | **76.9** | **3.7** | **79.8** | **4.0** | **39.3** | **18.1** | **71.9** |

<sup>1</sup>: from carpark entrance <sup>2</sup>: to entrance

Government gyms have the highest average compliance (84%), followed by university gyms (80%) and chain gyms (77%), with independent gyms having the lowest compliance (39%). Of the accessible parking component features, the length of the parking space and its visibility from the parking entrance are poorest; only 20% and 50% respectively passing the recommended requirements. The width and surfacing could also be improved.

Other studies on gym accessibility for PWD usually consider only one type of gym and only report on the number of accessible parking spaces. They confirm that while government-funded gyms generally have adequate provision (Dolbow & Figoni, 2015), accessible parking is still a barrier (Georgiou & Kaprinis, 2022; Reklaitiene et al., 2016).

#### 3.1.2 Main entrance

Four aspects of the main entrance were considered, namely the door width, entrance visibility, door operation and entrance threshold. A comparison of these with the recommendations in NZS4121: 2001 are presented in Table 4.

**Table 4.**
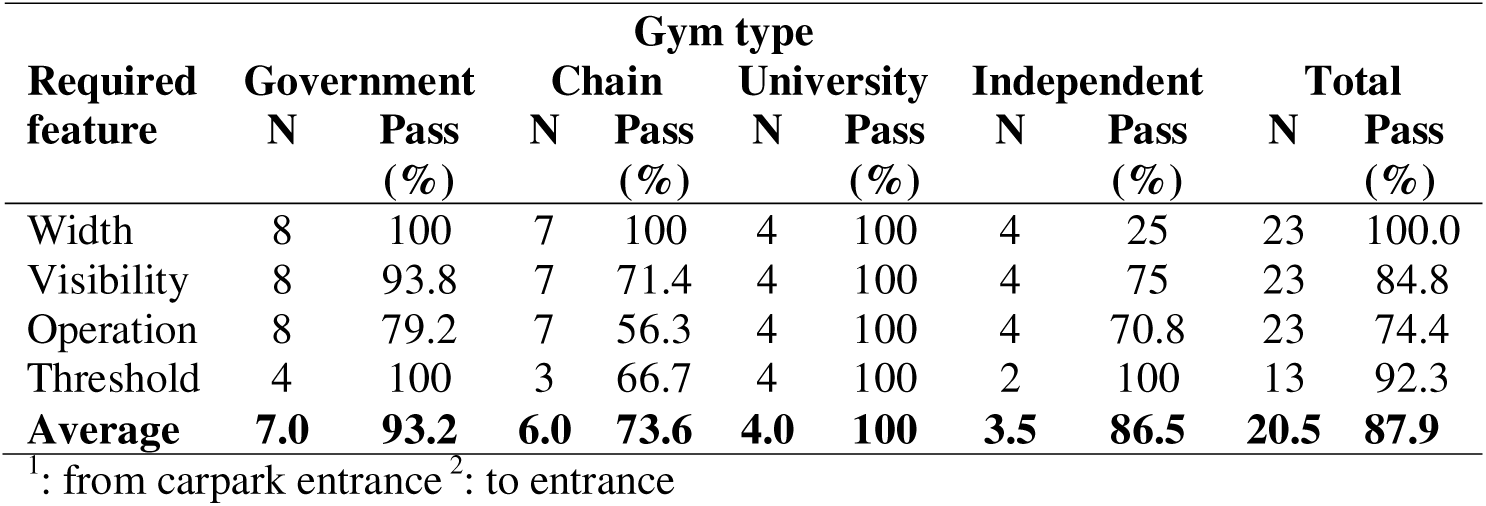
Audit results for the main entrance features in different types of gyms.

| Required feature | Gym type |  |  |  |  |  |  |  |  |  |
| --- | --- | --- | --- | --- | --- | --- | --- | --- | --- | --- |
|  | Government |  | Chain |  | University |  | Independent |  | Total |  |
|  | N | Pass (%) | N | Pass (%) | N | Pass (%) | N | Pass (%) | N | Pass (%) |
| Width | 8 | 100 | 7 | 100 | 4 | 100 | 4 | 25 | 23 | 100.0 |
| Visibility | 8 | 93.8 | 7 | 71.4 | 4 | 100 | 4 | 75 | 23 | 84.8 |
| Operation | 8 | 79.2 | 7 | 56.3 | 4 | 100 | 4 | 70.8 | 23 | 74.4 |
| Threshold | 4 | 100 | 3 | 66.7 | 4 | 100 | 2 | 100 | 13 | 92.3 |
| **Average** | **7.0** | **93.2** | **6.0** | **73.6** | **4.0** | **100** | **3.5** | **86.5** | **20.5** | **87.9** |

<sup>1</sup>: from carpark entrance <sup>2</sup>: to entrance

University and chain gym entrances have the highest average compliance (100% and 93% respectively). Chain gyms and independent gyms have the poorest entrances (73.6% and 86.5% respectively). Of the main entrance component features, the operation is the most problematic feature; automated doors closed too quickly or lacked markings showing the direction of opening while manual doors were heavy and hard to open. In addition, some main entrances were not clearly distinguishable from their surrounds making them difficult for people with low vision.

Previous research has found that gym entrances are an accessibility barrier for PWD (Reklaitiene et al., 2016).

#### 3.1.3 Internal features

Five aspects of the gym interior were considered, namely the internal doors (scored as an average of the clear open width, location of opening mechanism, colour contrast with surroundings, one-handed opening capability and force to open), passage (average of clearance and obstructions), service counter height, gym equipment clearance and water supply/drinking fountain height. A comparison of these with the recommendations in NZS4121: 2001 is presented in Table 5.

**Table 5.**
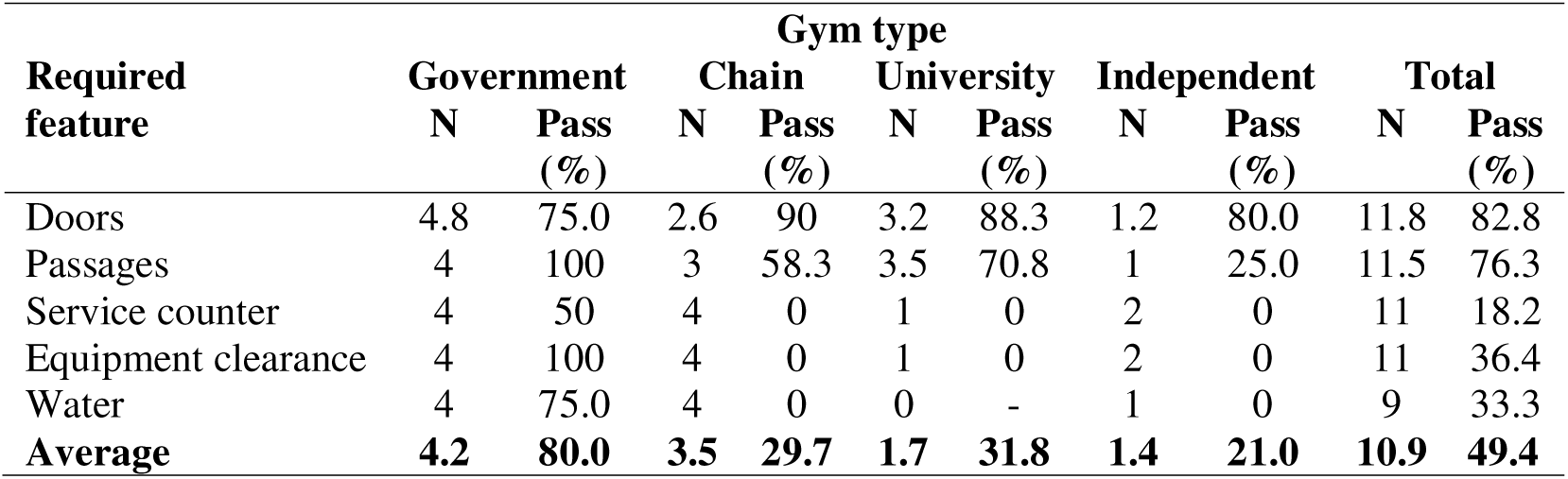
Audit results for the interior features in different types of gyms.

| Required feature | Gym type |  |  |  |  |  |  |  |  |  |
| --- | --- | --- | --- | --- | --- | --- | --- | --- | --- | --- |
|  | Government |  | Chain |  | University |  | Independent |  | Total |  |
|  | N | Pass (%) | N | Pass (%) | N | Pass (%) | N | Pass (%) | N | Pass (%) |
| Doors | 4.8 | 75.0 | 2.6 | 90 | 3.2 | 88.3 | 1.2 | 80.0 | 11.8 | 82.8 |
| Passages | 4 | 100 | 3 | 58.3 | 3.5 | 70.8 | 1 | 25.0 | 11.5 | 76.3 |
| Service counter | 4 | 50 | 4 | 0 | 1 | 0 | 2 | 0 | 11 | 18.2 |
| Equipment clearance | 4 | 100 | 4 | 0 | 1 | 0 | 2 | 0 | 11 | 36.4 |
| Water | 4 | 75.0 | 4 | 0 | 0 | - | 1 | 0 | 9 | 33.3 |
| **Average** | **4.2** | **80.0** | **3.5** | **29.7** | **1.7** | **31.8** | **1.4** | **21.0** | **10.9** | **49.4** |

Based on very limited data, gym interiors remain inaccessible for PWD. Government gyms were the most inclusive, passing most of the requirements for doors, passages, equipment clearance and drinking fountains but only half providing a lower service counter for people in wheelchairs. The latter is a common failure in all gym types. It is also important to recognise that people in wheelchairs need adequate space around equipment for transfer and should be able to reach drinking fountains. Similar barriers relating to doors, thresholds, equipment clearance, service counters and drinking fountains are reported in the literature (Mitchell et al., 2025a, b; Hansen et al., 2021; Calder et al., 2018)

#### 3.1.4 Accessible bathrooms

Eight aspects of the accessible bathrooms were considered, namely the location, signage, door clearance, floor area, toilet pan height, toilet handrail, hand basin and visibility of fittings. A comparison of these with the recommendations in NZS4121: 2001 are presented in Table 6.

**Table 6.**
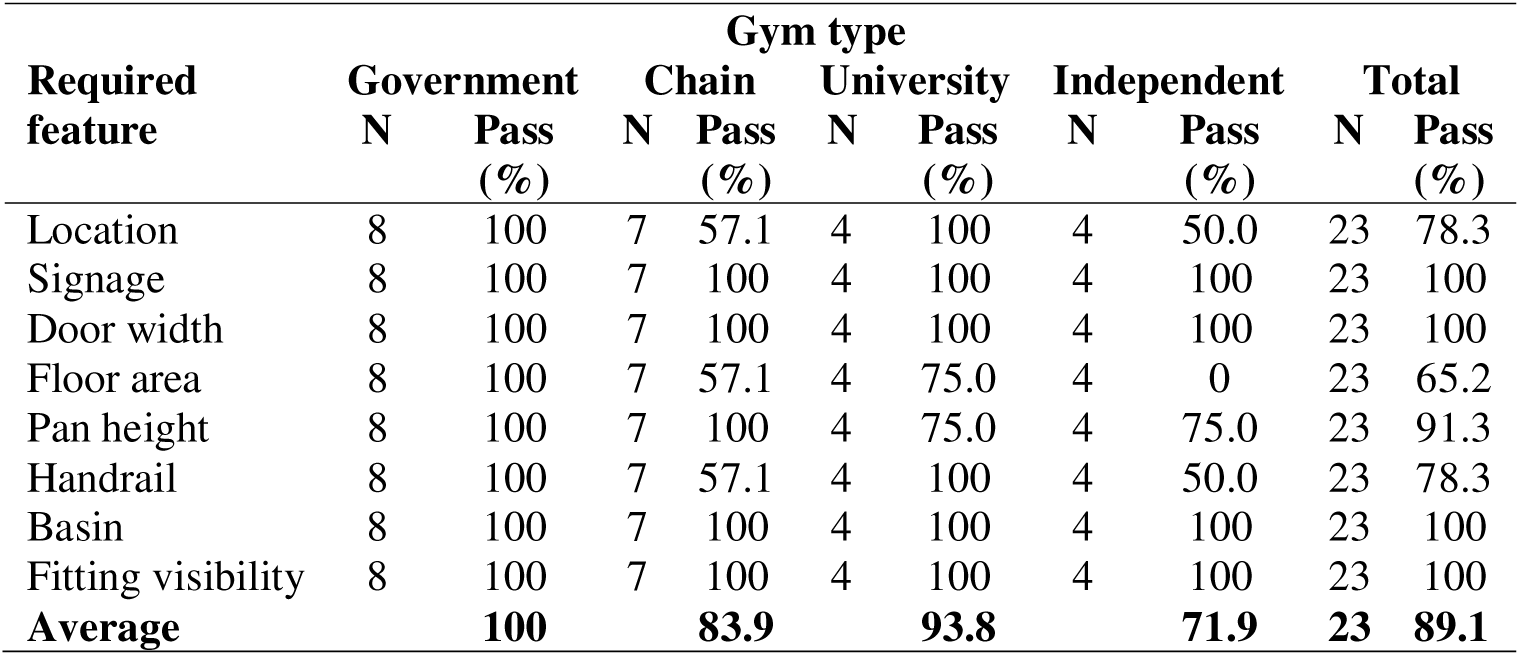
Audit results for the accessible bathroom features in different types of gyms.

The most accessible bathrooms are found in government gyms and university gyms. The weakest aspects of accessible bathrooms are inadequate floor space (particularly in independent gyms and chain gyms), not being located at/near the gym and not having an appropriate handrail at the toilet. Inadequate provision of accessible bathrooms and changing spaces is also reported in other studies (Dolbow & Figoni, 2015; Reklaitiene et al., 2016)

### 3.2 Interview findings

A summary of the responses to the open-ended questions is given in Table 7.

**Table 7.** Summary of responses to the open-ended interview questions, with number of responses, N.

| Question | N | Responses |
| --- | --- | --- |
| 1. What do you think a person with a disability is? | 16 | A person who has physical and/or mental problems. The disability can be hidden or visible. Anyone who needs help. |
|  | 1 | *“Someone who finds day-to-day life more difficult than the normal healthy person.”* |
| 2. Is access compliance and enforcement a priority? | 16 | 88% of respondents did not know, but those in larger gyms (government, university and chain) assumed that this was mandatory for the gym to operate. |
|  | 1 | University Facilities Manager: compliance requirements are clear and regular audits of all buildings ensure compliance. However, there are gaps in enforcement, particularly related to maintenance operations which might make certain areas temporarily inaccessible to PWD (eg. an alternative route not usable by someone in a wheelchair). |
| What accessibility features does the gym have for PWD? | 23 | Independent gyms have the basic features (access paths; an accessible toilet; handrails on stairs). Government gyms provide the most features, including ramps into pools; hoists (in changing rooms and the spa); changing facilities with full-size beds; automated doors; low counters; waterproof wheelchairs; hearing loops; emergency lanyards; elevators. |
| What accessibility barriers are there/what could be improved? | 23 | Stairs to upper viewing area - needs an elevator. Car parking for PWD could be better. Poolside disability changing rooms are hard to use for some people. Not all pools have ramp access. Malfunctioning assistive devices in accessible change rooms. Lights are too bright. It can be noisy. Customer service could be more inclusive. Provision of alarms in disabled spaces. |
| Have you had any training in handling PWD? | 16 | 81% of respondents have no training other than job experience. Regular, formal training is mandatory for government gyms. Chains have training that may include some information on PWD. Independent gyms rely on the owner being proactive about training. |
| What types of impairments have you encountered? | 16 | Mostly mobility impairments (people in wheelchairs, using crutches), some vision and hearing impairments, occasionally cognitive impairments. |
| What types of disability are best accommodated? What needs to improve? | 23 | Physical impairments, especially related to mobility. General recognition of the need to address a wider range of disability types, especially cognitive ones and to have some basic skills such as sign language. |
| Does the gym provide programs specifically for PWD? | 17 | 41% (mostly independent gyms) offer nothing beyond a personal trainer. The remainder offered mostly programs for older people. Government gyms offered specific programs eg. weekly 'Funky Movements' for PWD with caregivers. University gyms provided facilities for regular wheelchair sports and Special Olympics groups. |
| Are PWD included in the protocol for emergency evacuation? | 17 | 63% have no specific procedure designed for PWD beyond meeting the standard evacuation escape route requirements (clear path ending at a safe meeting place). Government gyms were best for PWD with staff trained to fetch assistive devices and help PWD evacuate and clicker alerts for visually impaired people. University and chain gyms provided some features eg. a voluntary log to record PWD presence with staff assistance in an evacuation; a fire alarm activated E-light system to alert visually impaired people. |

**Table 8.** Summary of average Likert scale responses for managers and staff.

| Question | N | Average |
| --- | --- | --- |
| Ranking scale: 1=Expert; 2=Very good; 3=Average; 4=Not much; 5=Nothing |  |  |
| 1. How much do you know about the challenges PWDs have when they try to access your gym | 17 | 2.50-2.91 |
| 2. How much do you know about the legal requirements for building accessibility features for PWDs? | 16 | 3.60-4.00 |
| Ranking scale: 1=Completely accessible; 2= Mostly accessible; 3=Not sure; 4=Not very accessible; 5=Not accessible at all |  |  |
| 3: How would you rate the overall accessibility of your gym for PWD) | 16 | 1.80-2.73 |
| Ranking scale: 1=Extremely important; 2=Quite important; 3=Not sure; 4=Not very important; 5=Not important at all |  |  |
| 4. How important is it to you that your building is accessible to all types of PWDs? | 16 | 1.00 |
| Ranking scale 1= very interested; 2=quite interested; 3=neutral; 4=quite uninterested; 5=completely uninterested |  |  |
| 5. How interested are you in receiving training on effectively interacting with and supporting PWD? | 16 | 1.80-1.92 |

The respondent’s opinion on the definition of a PWD were generally accurate, for example, “anyone who needs help”, but in one instance was ableist in distinguishing PWD from a “normal healthy person”. Most gym staff were ignorant about accessibility compliance requirements unless they engaged with it in their role as the facilities manager. The latter was aware that maintenance activities sometimes compromised accessibility, which has been identified as a common issue in other types of buildings (Zallio & Clarkson, 2021). Physical accessibility features and accessibility barriers were readily identified. However, staff acknowledged that support services were lacking, particularly those relating to the provision of PWD-specific exercise programs and proper staff training. In general staff wanted proper trained on interacting and supporting PWD. Similar findings are reported in the literature, namely that programs need to be more inclusive (Mitchell et al., 2025 a, b; Smith et al., 2021; Sharon-David et al., 2021) and that staff training needs to extend beyond basic awareness of PWD (McKenzie et al., 2025). Government gyms had the most comprehensive physical and social support systems while private gyms had the least. Lesch et al. (2025) report that for-profit gyms are less likely to have the financial resources to support inclusive accessibility and are generally more passive about the issue. All respondents thought that it was extremely important for gyms to be accessible to all types of PWD, although they may have felt morally obliged to express this opinion.

## 4. Discussion and Conclusion

The case study of New Zealand gyms, ranging from large government-funded community facilities to small, privately-owned businesses highlights significant accessibility barriers related to physical infrastructure, sensory accommodations, and service provisions for PWD. Quantitative accessibility audits based on the requirements of NZS4121:2001 were used to assess the compliance of accessible parking, entrances, interiors and bathrooms. The most accessible aspects include parking spaces, footpaths, entrance doors and bathrooms with features such as clear signage, well-designed ramps, and evacuation protocols demonstrating best practice inclusivity. Improvements in clear space, service counters, lifts, tactile and auditory communication are needed. Interviews with staff and managers provided qualitative data on operational practice and awareness of inclusivity. Community gyms and university gyms are the most inclusive and their staff have the most comprehensive experience in interacting with people with different disabilities. Small private gym owners know little about accessibility barriers and have financial constraints that limit their actions to address accessibility barriers. The main recommendations for improving gym accessibility for people of all abilities include more extensive staff training and regular maintenance of the facility’s accessible features. The training needs to extend beyond basic disability awareness training and include training on meeting the exercise and support needs of PWD in the gym environment. Gyms also need to offer more exercise programs targeted to different types of disability and encourage attendance by support carers. These strategies will help reduce some of the barriers for those PWD who already go to the gym. However, there remains the problem of getting PWD to begin going to a gym. A review of the literature highlighted the poor participation in exercise by PWD. Gym participation can be improved by educating PWD on the health benefits of regular exercise (and the health risks of being physically inactive) and by removing the gym membership cost barrier. The involvement of healthcare professionals is critical; they are uniquely placed to tell their PWD clients about the health benefits and risks, to prescribe gym membership so that it can be subsidised by social programs and to monitor the client’s health as a proxy for gym attendance. There is a real cost benefit from this strategy: the cost of providing physical exercise far outweighs the future health costs of people who are inactive. There is an additional benefit to the quality of life of PWD from the increased participation in exercise and society.

These findings are significant. Accessibility is a fundamental human right enhancing social inclusion, independence, and well-being of PWD. The research emphasises the importance of gyms adopting a more inclusive and proactive approach to accessibility, ensuring that facilities are welcoming and accommodating to all users. It also emphasises the need for proactive participation of healthcare professionals and governments in encouraging PWD to exercise regularly. The research impacts other stakeholders, including gym staff, facility managers, policymakers, and advocacy groups. Gym staff and facility managers can apply the study’s recommendations to improve the PWD experience and to implement practical, cost-effective solutions that improve accessibility. Policymakers can use these findings to reinforce regulatory frameworks and enforcement measures, ensuring that accessibility standards are not merely suggestions but mandatory regulations. Finally, advocacy groups can promote the recommendations and hold relevant parties accountable.

A large proportion of our population lives with disabilities and has a heightened risk of secondary physical health problems and psychological health problems. Regular exercise in gyms can reduce this risk and ultimately lead to a better quality of life.

## Data Availability

All data produced in the present work are contained in the manuscript

## Acknowledgements

The authors supervised three students in Massey University’s School of Built Environment who collected the underlying data as part of their required research component in the Master of Construction degree. The authors acknowledge the student’s efforts.

